# Breastmilk; a source of SARS-CoV-2 specific IgA antibodies

**DOI:** 10.1101/2020.08.18.20176743

**Authors:** Britt J. van Keulen, Michelle Romijn, Albert Bondt, Kelly A. Dingess, Eva Kontopodi, Karlijn van der Straten, Maurits A. den Boer, Berend J. Bosch, Philip J.M. Brouwer, Christianne J.M. de Groot, Max Hoek, Wentao Li, Dasja Pajkrt, Rogier W. Sanders, Anne Schoonderwoerd, Sem Tamara, Rian A.H. Timmermans, Gestur Vidarsson, Koert J. Stittelaar, Theo T. Rispens, Kasper A. Hettinga, Marit J. van Gils, Albert J.R. Heck, Johannes B. van Goudoever

## Abstract

**Background:** Since the outbreak of COVID-19, many put their hopes in the rapid development of effective immunizations. For now patient isolation, physical distancing and good hygiene are the sole measures for prevention. Processed breast milk with antibodies against SaRS-CoV-2 may serve as additional protection. We aimed to determine the presence and neutralization capacity of antibodies against SaRS-CoV-2 in breastmilk of mothers who have recovered from COVID-19.

**Methods:** This prospective case control study included lactating mothers, recovered from (suspected) COVID-19 and healthy controls. Serum and breastmilk was collected. To assess the presence of antibodies in breastmilk and serum, we used multiple complementary assays, namely ELISA with the SARS-CoV-2 spike protein, SARS-CoV-2 receptor binding domain (RBD) and with the SARS-CoV-2 nucleocapsid (N) protein for IgG and bridging ELISA with the SARS-CoV-2 RBD and N protein for total Ig. To assess the effect of pasteurization breastmilk was exposed to Holder Pasteurization and High Pressure Pasteurization.

**Results:** Breastmilk contained antibodies against SARS-CoV-2 using any of the assays in 24 out of 29 (83%) proven cases, in six out of nine (67%) suspected cases and in none of the 13 controls. *In vitro* neutralization of SARS-CoV-2 clinical isolate virus strain was successful in a subset of serum (13%) and milk samples (26%). Although after pasteurization of the milk SARS-CoV-2 antibodies were detected with both methods of pasteurization, virus neutralizing capacity of those antibodies was only retained with the HPP approach.

**Conclusion:** Breastmilk of mothers who recovered from COVID-19 contains significant amounts of IgA against SARS-CoV-2, both before and after pasteurization.

**Key Points:** *Question:* Does breastmilk of mothers who have recovered from coronavirus disease 2019 (COVID-19) contain antibodies against severe acute respiratory syndrome coronavirus 2 (SARS-CoV-2)?

*Findings:* We provide multiple lines of evidence on the presence of a variety of antibodies against SARS-CoV-2, with no such antibodies present in the controls. These antibodies are capable of neutralizing a clinical isolate of SARS-CoV-2 *in vitro*. We furthermore show that high pressure pasteurization hardly affects antibody levels and efficacy.

*Meaning:* Breastmilk, obtained from mothers who have recovered from COVID-19, may serve as a safe and widely applicable preventive strategy for vulnerable high risk populations

## Introduction

The severe acute respiratory syndrome coronavirus 2 (SARS-CoV-2) outbreak, which was first reported in December 2019, has had an enormous global impact. SARS-CoV-2 can cause coronavirus disease 2019 (COVID-19) with the number of confirmed cases over 20 million, and approximately 750,000 deaths globally as of August 12^th^ 2020.

In response to the pandemic, many countries have had to introduce drastic lockdowns to enforce physical separation, which are affecting economies worldwide, while also imposing a huge psychological burden to specific groups such as elderly. On a personal level, general preventive measures like protective materials, physical distancing and frequent hand washing, have shown to be effective, but the pandemic necessitates rapid development of effective vaccines as prevention. Despite the ongoing development of over 100 vaccine candidates, of which already 10 are in clinical trials, an effective and widely available vaccine is not yet available [1].

When looking at preventive strategies, it is interesting to note that, in infants, breastfeeding is associated with a 30% reduction in respiratory infections when compared to formula feeding [2, 3]. It is generally accepted that this protective effect is due to the human immune components in breastmilk, such as specific antibodies as of which secretory immunoglobulin A (sIgA) is the most abundant. SIgA represents our first line of defense as it acts directly at mucosal surfaces [4]. SIgA inhibits microbial binding to host receptors of intestinal epithelial cells, entrapping pathogenic microorganisms within the mucus and enhances ciliary activities, thus eliminating invading pathogens [5–7]. Through this mechanism, breast milk sIgA may provide protection against entry of the SARS-CoV-2 virus in the airway mucosal surface.

The structural proteins of SARS-CoV-2 include the spike (S), nucleocapsid (N), membrane, and envelope proteins (**Figure 1**). The S1 subunit of the S protein contains the receptor binding domain (RBD) which facilitates angiotensin-converting enzyme 2 (ACE2) receptor mediated virus attachment, while the S2 subunit of the S protein promotes membrane fusion to initiate the infection of host cells. The N protein encapsulates viral RNA and is necessary for viral transcription and replication [8]. The human immune system will, when infected by SARS-CoV-2, generate antibodies against one or more of these viral proteins, whereby variability may exist in the immunoglobulin class preferentially made (e.g. IgG, IgA, IgM). The titers of antibodies are individual specific, but also determined by the severity of the infection and the time that has passed since the onset of the infection. Therefore, it is recommended to use complementary assays to determine SARS-CoV-2 antibody titers.

**Figure 1:**
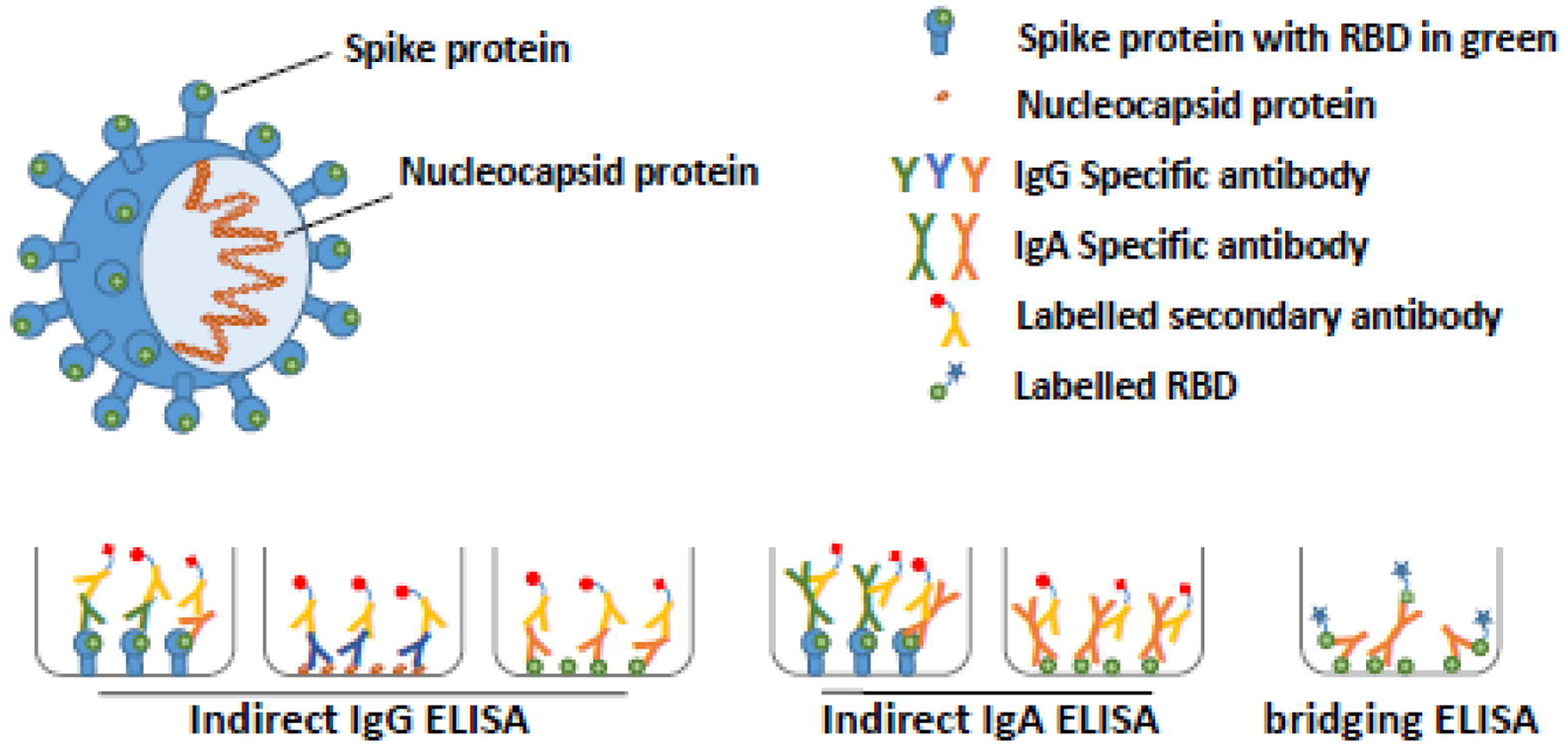
Schematic representation of SARS-CoV-2 and the different ELISA assays used to detect SARS-CoV-2-reactive antibodies. The spike (including the receptor binding domain (RBD)) and nucleocapsid proteins of SARS-CoV-2 are depicted in the context of the virus. SARS-CoV-2 specific antibodies were detected using multiple complementary ELISA assays. The indirect ELISA assays using S, RBD or N were used to detect IgG or IgA specific-antibodies (green, blue or orange, respectively) and the bridging ELISA assay was used to detect total Ig against the RBD.

There is strong evidence that antibodies, especially of the IgA class, against several respiratory infections, such as influenza, are secreted into breastmilk [9, 10]. Pre-print data indicates that 15-30 days following the onset of symptoms, antibodies against the SARS-CoV-2 RBD may be present in breastmilk [11]. These data form the basis of our hypothesis that an array of SARS-CoV-2-reactive milk antibodies may be present in breastmilk of mothers who have recovered from COVID-19. Their milk could potentially be used clinically to reduce the incidence of SARS-CoV-2 infections, by neutralizing the virus in the airway mucosa, although many steps need to be taken before such an approach can be implemented. These include, amongst others, pasteurization of the milk. Breast milk may contain pathogens, and therefore pasteurization is required prior to use, especially in a vulnerable population [12]. Holder pasteurization (HoP), a heat treatment at 62.5°C for 30 min, is currently the standard pasteurization method [13]. Although HoP effectively inactivates microbial contaminants, it concomitantly reduces the activity of some important bioactive milk components [13, 14]. For this purpose, alternative methods to HoP, such as high pressure pasteurization (HPP), are currently being investigated [15, 16]. Our aim was thus to evaluate the level of antibodies in untreated breast milk, but also after thermal (HoP) and non-thermal (HPP) pasteurization and to determine the efficacy of virus neutralization of serum and untreated, and pasteurized milk.

## Methods

### Study population

This prospective case control study included lactating women with a confirmed or high probability of COVID-19. Lactating women who recovered from a proven COVID-19 infection were recruited by an online recruitment letter. A confirmed infection was defined as a positive SARS-CoV-2 PCR from a nasal-pharyngeal swab. Subjects were classified in the suspected COVID-19 group in the event of a confirmed infection with SARS-CoV-2 in the household and the lactating woman developed COVID-19 symptoms as well. A control group of healthy lactating women were recruited from the Amsterdam UMC if they met the following criteria: lactating women who delivered at Amsterdam UMC with a negative SARS-CoV-2 PCR from a nasal-pharyngeal swab during delivery, and without symptoms of COVID-19.

Ethical approval was obtained from the Medical Ethics Committee of the Amsterdam UMC, location VUmc and written informed consent was obtained from all participants.

### Data collection

All participants were requested to collect 100 ml of breastmilk from an entire breast in specially provided bottles and to store the bottle in their freezer until collected by study staff during a home visit. Subsequently, the samples were stored at −20°C. During the home visit, maternal serum was collected by a trained phlebotomist.

## Laboratory analysis

### Evaluation of antibodies in the breastmilk and serum

To assess the variety and variability of antibodies potentially present in the subjects, we decided to use multiple complementary assays (**Figure 1**), namely ELISA with the SARS-CoV-2 spike protein (breastmilk and serum; for IgA and IgG, respectively), ELISA with the SARS-CoV-2 receptor binding domain (RBD) and with the SARS-CoV-2 nucleocapsid (N) protein for IgG (serum only) and bridging ELISA with the SARS-CoV-2 RBD and N protein for total Ig (breastmilk and serum).

#### ELISA with the SARS-CoV-2 Spike protein

Soluble perfusion-stabilized S-protein of SARS-CoV-2 using stabilization strategies were generated as previously described [17]. This protein was immobilized on a 96-well plate (Greiner) at 5 µg/mL in 0.1 M NaHCO3 overnight, followed by a one-hour blocking step with casein (Thermo Scientific). Breastmilk was diluted 1:5 and serum was diluted 1:100 in casein and incubated on the S-protein coated plates for 2 hours to allow binding. Antibody binding was measured using 1:3000 diluted HRP-labeled goat anti-human IgG (Jackson, Immunoresearch) in casein for the serum samples and 1:3000 diluted HRP-labeled goat anti-human IgA (Biolegend) in casein for the breastmilk samples.

#### ELISA with the SARS-CoV-2 total RBD in breastmilk and RBD-Ab, RBD-IgG and Nucleocapsid protein in serum

Antibodies against RBD protein were measured in a total antibody ELISA and IgG ELISA essentially as described before [18]. Briefly, for total antibodies, samples were incubated (1:10 for serum, undiluted for breast milk) on plates coated with RBD protein and antibodies detected using biotinylated RBD protein. For IgG, serum samples were diluted 1:100 and incubated on RBD-coated plated, followed by detection of IgG antibodies using a mouse monoclonal antihuman IgG antibody. Total antibodies against NP in serum were measured in 1:10 diluted serum on NP-coated plates followed by detection using biotinylated NP protein.

### Effect of antibodies on virus replication

In order to assess if breastmilk with SARS-CoV-2 possess virus neutralizing activity, *in vitro* neutralizing assays were conducted using a SARS-CoV-2 clinical isolate strain, which was kindly provided by Christian Drosten, Charité-Universitätsmedizin, Berlin, Germany (BetaCoV/Munich/BavPat1/2020), performed under biosafety level 3+ conditions. In brief, 60 μL of SARS-CoV-2 working dilution containing approximately 200 TCID50/well was mixed with 60 μL of serially 2-fold dilutions of heat-inactivated serum or milk, in triplicates and incubated for 60 minutes at 37°C to allow for neutralization of the virus. Subsequently 100 μL of these virus/antibody mixtures were added to confluent VERO E6 cell monolayers (ATCC; CRL-1586) and incubated at 37°C for four to six days. The virus working dilution and the original virus stock were titrated in a parallel plate and served as positive virus controls in each assay run. After incubation at 37°C 20 μL of a WST-8 Cell Counting Kit-8 (CCK-8) solution (Sigma-Aldrich; 96992) was added to each well of the plate, followed by an incubation for three hours at room temperature. The absorbance at 450 nm was measured using microplate reader (Biotek Synergy H1). The Reed and Muench method was used to determine the 50% end-point titer of the sample, as well as the virus titer (stock and back titration).

### Evaluation of the effect of pasteurization of breastmilk on SARS-CoV-2 antibodies

We used two methods of pasteurization and after these treatments evaluated the amount of SARS-CoV-2 antibodies and neutralizing capacity of breastmilk. During Holder pasteurization (HoP), currently the standard method for pasteurization, breastmilk is pasteurized at 62.5 °C for 30 min. An alternative to HoP pasteurization is high pressure pasteurization (HPP), which inactivates vegetative (pathogenic) micro-organisms, yeasts, molds and viruses, with no heat-induced damage [19].

Samples were stored frozen at –20 °C and thawed overnight in a refrigerator (7 °C), prior to being transferred into sterile pouches that were double packed and treated at a pilot-scale high-pressure unit with water at ambient temperature [20]. We applied a hydrostatic pressure of 500 MPa for 5 minutes. All samples were stored at −20°C directly after treatment.

### Monitoring IgA clone diversity in breastmilk before and after pasteurization by mass spectrometry

In addition to the classical antibody detection assays, we used a novel mass spectrometry (MS) method to examine the IgA clonal diversity in breastmilk. We examined the IgA clones in unpasteurized breastmilk, and after the two different pasteurization techniques. The antigen binding fragments (Fab) were proteolytically released from the captured IgAs, and the resulting Fab fragments (45-50 kDa) of individual clones were profiled using MS. The abundance of each unique detected clone could be determined, and thus for each clone the effect of the two different pasteurization techniques could be monitored.

## Statistical analysis

Statistical analysis was performed with IBM SPSS Statistics for Windows, version 26 (IBM Corp., Amonk, N.Y., USA). Patient characteristics and COVID-19 symptoms were expressed as mean with standard deviation (SD) or median with interquartile range (IQR) depending on their distribution.

## Results

Our prospective case control study included 38 lactating women with a confirmed or high probability of COVID-19 and 13 healthy controls. Three of the subjects with a confirmed infection were admitted to the hospital. Samples were obtained at different time intervals from the onset of clinical symptoms, as shown in **Table 1**.

**Table 1.** Patient characteristics of the lactating women with a confirmed or highly probable COVID-19 and controls. NPS = nasal-pharyngeal swab, SD = standard deviation, IQR = interquartile range, NA = not applicable.

| Characteristics | Confirmed COVID-19<br>N=29 |  | Suspected COVID-19<br>N=9 |  | Controls<br>N=13 |
| --- | --- | --- | --- | --- | --- |
| Gestational age – weeks median (IQR) | 39.7 (38.5, 40.7) |  | 38.8 (36.8, 40.4) |  | 40.7 (39.6, 41.1) |
| Age of child – weeks median (IQR) | 28.9 (12.1, 39.5) |  | 12.6 (8.4, 40.7) |  | 6.1 (4.3, 7.4) |
| Age of mother – years mean (SD) | 31.1 (3.1) |  | 30.3 (4.1) |  | 33.2 (3.3) |
| Time between start of clinical symptoms and collection of breast milk – weeks mean (SD) | 5.9 (2.6) |  | 5.7 (2.1) |  | NA |
| **Symptoms and duration in days** | **N. (%)** | **Median (IQR)** | **N. (%)** | **Median (IQR)** |  |
| Fever >37.5 °C | 21 (72%) | 3 (1, 5) | 6 (67%) | 1 (0, 4) | NA |
| Cold | 24 (83%) | 12 (5, 20) | 4 (44%) | 7 (4, 54) | NA |
| Cough | 21 (72%) | 14 (5, 28) | 5 (56%) | 6 (3, 12) | NA |
| Sore throat | 21 (72%) | 6 (4, 14) | 5 (56%) | 6 (3, 12) | NA |
| Tachypnea | 5 (17%) | 11 (4, 14) | 2 (22%) | NA | NA |
| Dyspnea – | 14 (38%) | 7 (3, 28) | 1 (11%) | NA | NA |
| Stomachache | 5 (17%) | 2 (1, 9) | 2 (22%) | NA | NA |
| Nausea | 5 (17%) | 3 (2.5, 14) | 1 (11%) | NA | NA |
| Vomiting | 2 (7%) | NA | 0 | NA | NA |
| Diarrhea | 5 (17%) | 2 (1, 25) | 3 (33%) | NA | NA |
| Headache | 24 (83%) | 5 (2, 12) | 8 (89%) | 7 (4, 14) | NA |
| Photophobia | 2 (7%) | NA | 0 | NA | NA |
| Anosmia | 18 (62%) | 20 (13) | 6 (67%) | 9 (4, 22) | NA |
| Ageusia | 17 (59%) | 19 (13) | 4 (44%) | 14 (11, 25) | NA |
| Fatigue | 24 (83%) | 20 (16) | 8 (89%) | 10 (4, 36) | NA |
| Anorexia | 10 (34%) | 12 (8, 21) | 3 (33%) | NA | NA |
| Hospital admission | 3 (10%) | NA | 0 | NA | NA |
| Non-invasive respiratory support (O2) during admission | 2 (7%) | NA | 0 | NA | NA |

### SARS-CoV-2 antibodies in breastmilk and serum

Breastmilk contained antibodies against the SARS-CoV-2 virus, using any of the assays, in 24 out of 29 (83%) proven cases and in six out of nine (67%) of the suspected cases (**Figure 2**). None of the 13 controls expressed SARS-CoV-2-reactive antibodies of any kind in breastmilk. A large variability in antibody levels was found in the milk samples of the different subjects. Both the assay assessing IgA response against the S protein and the assay detecting the total Ig response against RBD showed a variable pattern in antibody type and the SARS-CoV-2 protein it recognized (**data in the supplemental eTable 1**).

**Figure 2.**
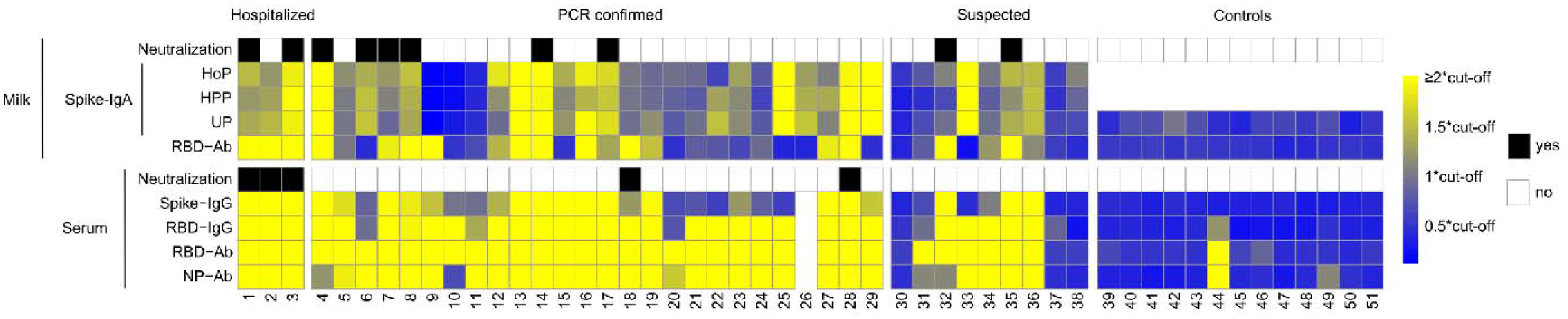
Multiple assay assessment of SARS-CoV-2 antibody levels in breastmilk and serum. Colors from blue to yellow indicate increasing levels of antibodies in milk and serum, relative to the cut-off values of the respective assays, with all levels over 2 times the cut-off being bright yellow, as indicated by the color scale. Neutralization by milk (either unpasteurized and/or high pressure pasteurized) or serum indicated as either positive (black) or negative (white). Ab; total Ig.

Secretion of antibodies may well be dependent on the time that has passed since the onset of the disease. By using a cross sectional sampling design, we were able to show that even up to 13 weeks from onset, detectable antibody levels were found (**supplemental eFigure 1**). While over 80% of the milk samples in the PCR proven cases contained antibodies, all of their blood samples showed a positive response in at least one of the four assays.

### SARS-CoV-2 virus neutralization

Next, we aimed to assess if breastmilk that contained antibodies against SARS-CoV-2 was able to reduce virus replication. Neutralization of the SARS-CoV-2 virus in this model was successful in a subset of serum (13%) and milk samples (26%), especially when these had relative high antibody (IgG in serum and IgA in milk, respectively) levels (**Figure 2 and Supplemental eFigure 2**). In two of the subjects, both serum as well as unpasteurized and HPP treated breastmilk was able to neutralize the SARS-CoV-2 virus. Both subjects (sample numbers 1 and 3 in **Figure 2**) had been admitted to the hospital. We could, however, not detect a linear correlation between milk antibody levels and the virus neutralization capacity (**Supplemental eFigure 2)**.

### Pasteurization of breastmilk

Studying IgA S protein before and after the two types of pasteurization, we did not observe a difference in their levels (**Figure 3a**)The virus neutralizing capacity of those milk samples was, however, only retained with the HPP approach, and not with HoP (**Supplemental eFigure 2**), indicating that heat treatment may have caused a loss-of-function even though this did not result in reduced antibody levels according to ELISA. This shows the importance of testing the functionality of the milk in a neutralization assay on top of the analysis of the antibody levels.

**Figure 3a:**
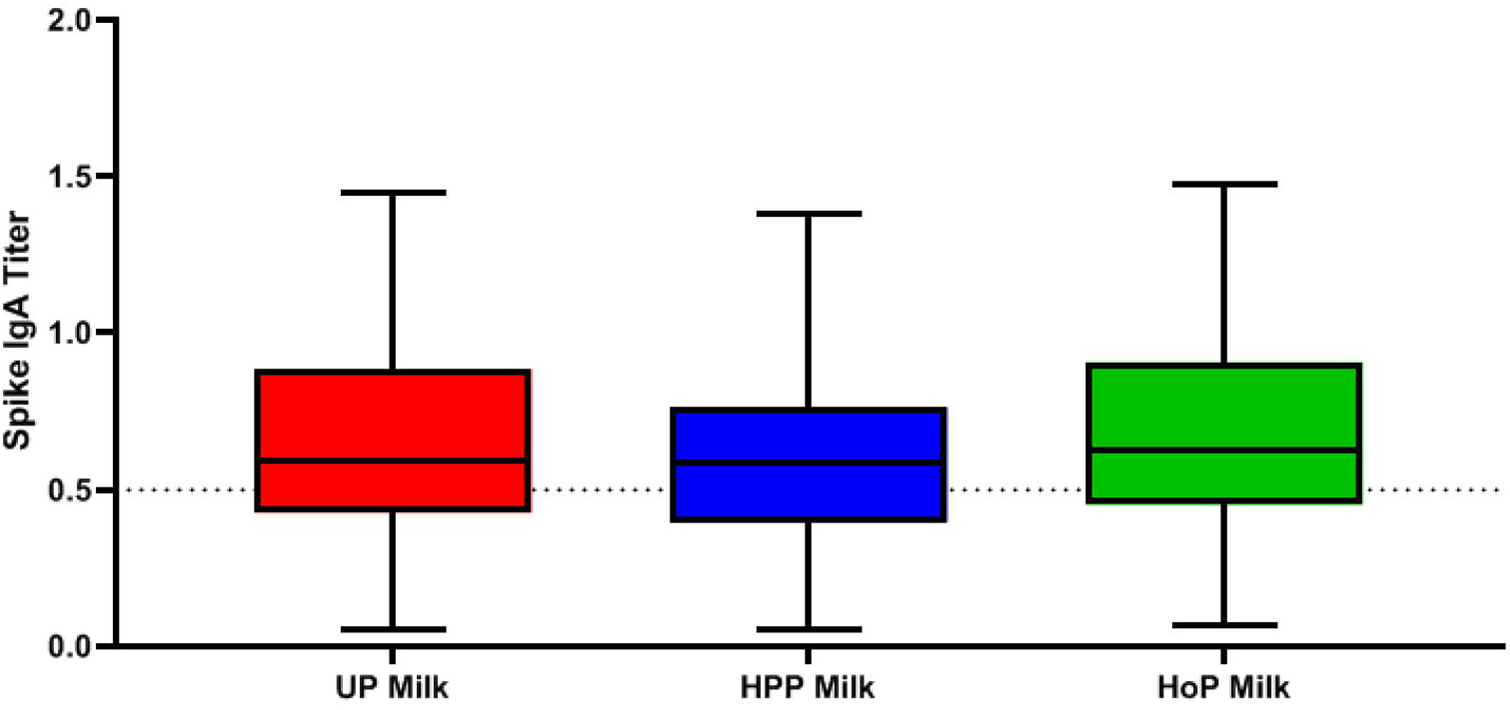
Spike IgA titers in unpasteurized (UP) milk, high pressure pasteurized (HPP) milk and holder pasteurized (HoP) milk.

**Figure 3b:**
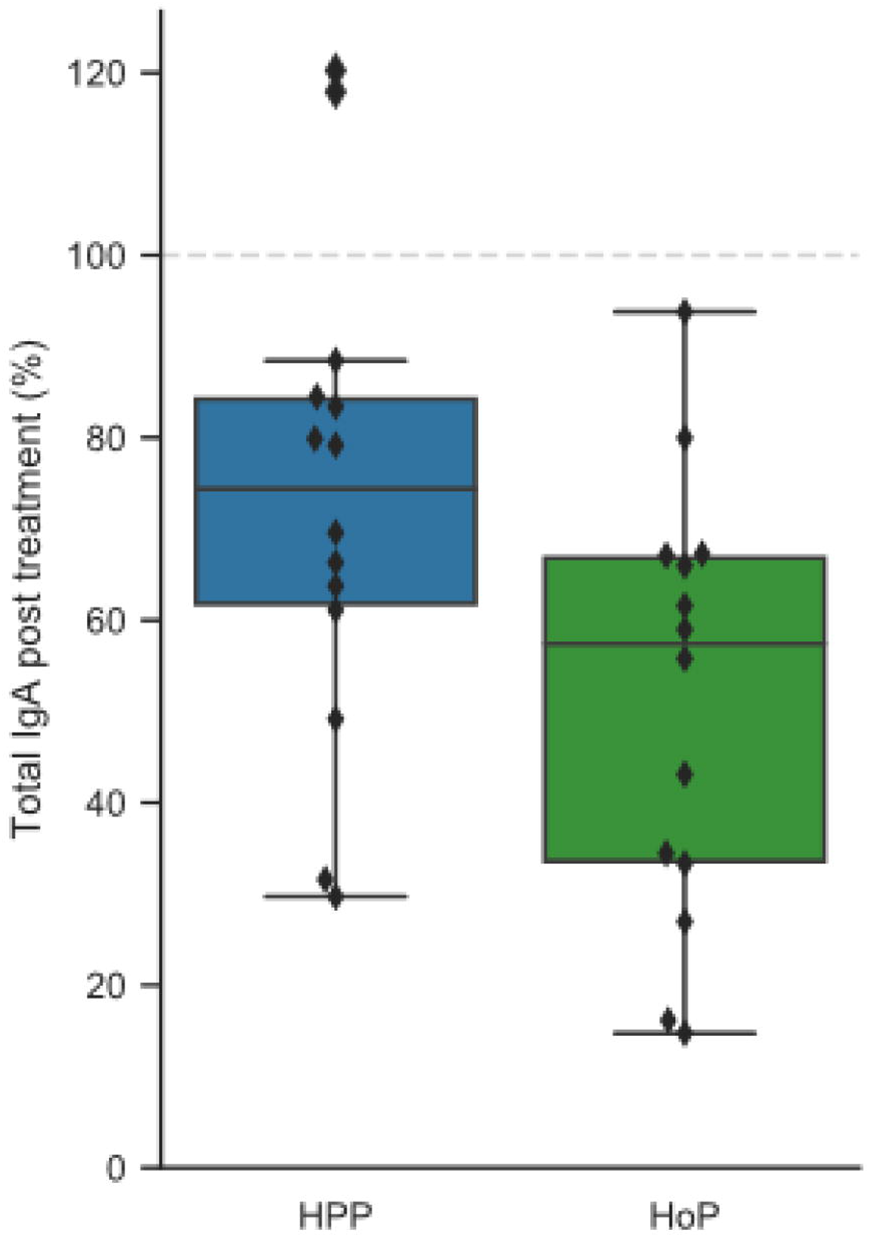
IgA retention according to LC/MS profiles following HoP and HPP expressed as the concentration of IgA relative to raw breastmilk.

### Monitoring IgA clone diversity in breastmilk before and after pasteurization by mass spectrometry

**Figure 4** shows the variability of the IgA profiles of three different subjects. Clearly, the IgA profiles are highly individual specific. The pasteurization effect is very limited in all cases and seems to affect all clones equally.

**Figure 4.**
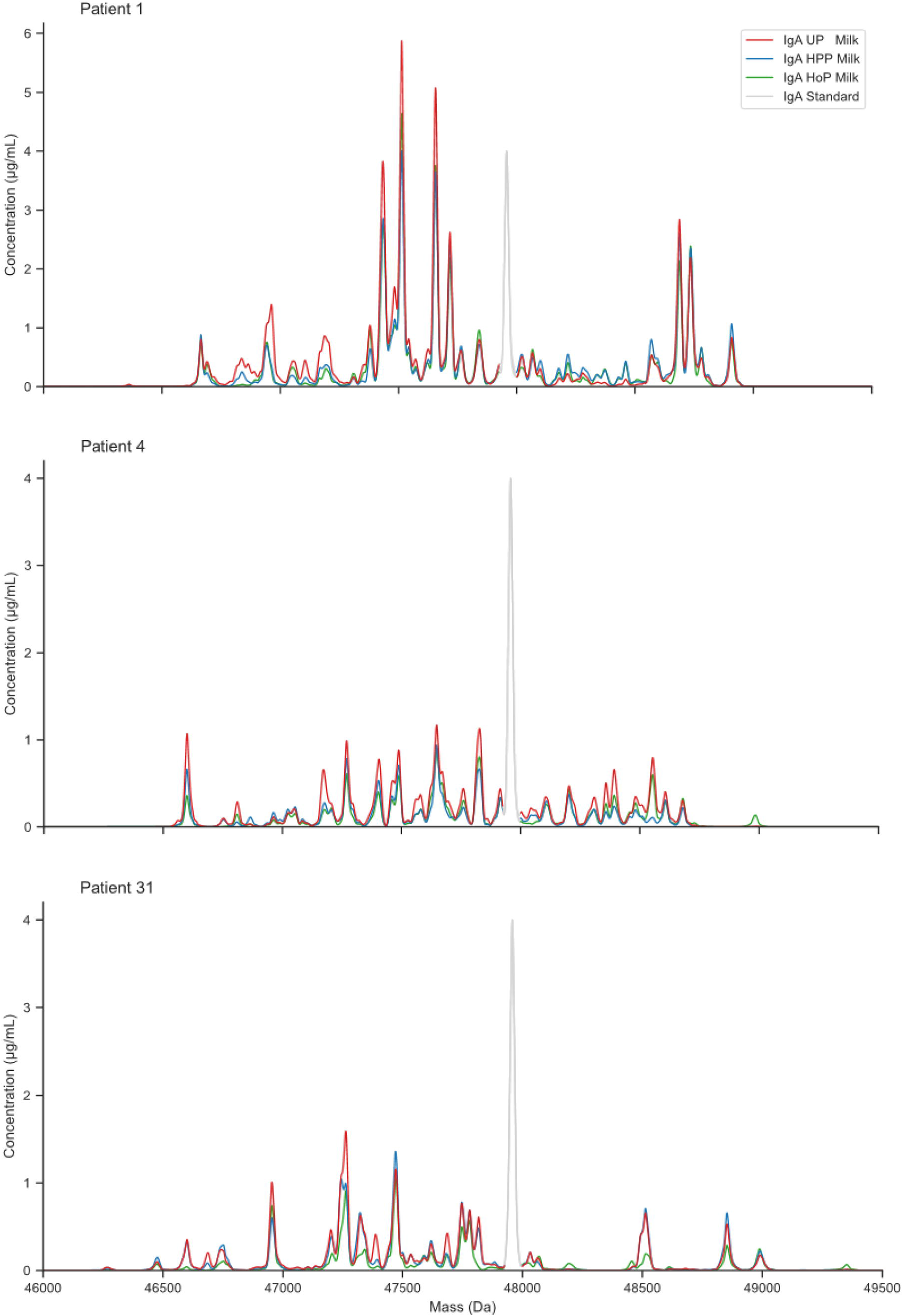
Exemplary Fab profiles of IgA clones present in breastmilk of individual subjects. Each individual peak represents a different clone with a distinct mass (Da) and abundance. The grey peak represents the signal of the spiked-in recombinant IgA standard used for quantification.

Following HoP and HPP, the abundance of each clone remained largely unaffected, although on average some loss was observed as summarized in **Figure 3b**. The reduction in overall IgA concentration was greater in HoP- than HPP-treated milk.

## Discussion

We demonstrate that breastmilk of mothers who recovered from COVID 19 contains significant amounts of IgA against SARS-CoV-2, both before and after HoP and HPP pasteurization. In addition, SARS-CoV-2 directed IgA levels in breastmilk are present over a long period of time (for at least 13 weeks following the onset of COVID-19 symptoms). We also show that serum and milk samples from some donors had neutralizing capacity against a SARS-CoV-2 clinical isolate *in vitro* resulting in significant inhibition of virus propagation. In cases where high antibody levels were present, both raw and HPP-treated milk were able to neutralize the virus, whereas HoP-treated milk was not.

Our data clearly indicate strong variability in individual antibody levels. Some subjects show a more N protein or RBD directed antibody response and some exhibit a stronger IgA or IgG response. The presence and abundance of SARS-CoV-2 specific antibodies is known to be variable, for instance, the IgG response to the N protein is believed to occur earlier than the response to the S protein, but the titers are lower to the N protein compared to the S protein [21]. Also, in general IgM and IgA-class responses often occur earlier following disease onset, while IgG responses occur later and seem to be longer lasting [22]. Together, these results imply that testing for the presence of antibodies should not be directed against a single protein or focus on a single antibody class, but that different classes and proteins should be targeted to obtain the most reliable information.

Human milk banks around the world use HoP as a way to provide donor milk to preterm and sick, term infants. However, HoP is known to affect the immune protection provided by breastmilk, due to the high heat load to which the milk is exposed. One promising alternative to HoP is HPP. HPP is already widely used in food industries as a non-thermal food preservation method that provides microbiologically safe products, while at the same time reducing the heat-induced damage of regular thermal pasteurization. Recent studies on HPP of donor breastmilk indicate that this method is capable of retaining significantly higher levels of antibodies when compared to HoP, while at the same time successfully eliminating microbes and viruses such as HIV and CMV [15, 16, 23–25]. Our data indicate that HPP would be a more suitable method to make breastmilk safe than thermal pasteurization, as indicated by the retention of functional IgAs in breastmilk, and the retained neutralization towards the clinical SARS-CoV-2 isolate.

This study has several strengths. First, we were able to collect clinical data, blood and human milk from almost all participants. Second, by using different methods to measure antibodies in serum and human milk we created robust data, capturing the variation in antibody responses. In addition, we included women with a different time window between the COVID-19 infection and time of sampling, and were therefore able to investigate the dynamics of the antibody levels over time. Third, by using virus neutralizing as functional readout, we were able to draw conclusions on the effectiveness of the found antibodies against the virus.

Breastmilk is known to be a safe product that can be used for preventive strategies, especially compared to pharmaceutical interventions (either medication or vaccination), and no detrimental side effects are to be expected from its intake. The virus itself is not present in breastmilk and by pasteurizing the milk it is possible to provide a safe product. Furthermore, breastmilk can be applied globally, and is readily available, also in the low income countries. However, using breast milk as a preventive strategy requires ample availability of breast milk from COVID 19 recovered women. While rates of seroprevalence of anti-SARS-CoV-2 IgG antibodies in the general population varies widely, “milkprevalence” rates in pregnant and lactating women are not known and might differ substantially from region to region.

The possibility to purify sIgA from breast milk from mothers who have recovered from COVID 19 opens also possibilities, but again implies availability of sufficient seroconverted milk.. Vulnerable populations such as front-line health-care workers and elderly may be prioritized for randomized controlled trials to establish efficacy of such preventive strategies. Even though we are still far away from clinical application, efforts should be undertaken to investigate the possibility of using breastmilk as a preventive strategy against SARS-CoV-2 infection and subsequent spread.

## Conclusions

Breastmilk of mothers who were previously infected with SARS-CoV-2 contained significant amounts of IgA against SARS-CoV-2, both before and after two modes of pasteurization, and also after a long period post-infection. Milk samples of several donors had neutralization capacity against a clinical isolate of SARS-CoV-2, also after non-thermal pasteurization.

## Data Availability

The authors confirm that the data supporting the findings of this study are available within the article [and/or] its supplementary materials. The data that support the findings of this study are available from the corresponding author upon reasonable request.

## Acknowledgements

Mirjam Damen and Arjan Barendregt (UU) are acknowledged for their assistance in the mass spectrometric analysis of the IgA clones. The Viroclinics Xplore analytical team is acknowledged for organizing and performing the virus neutralization assay.

AJRH acknowledges the Netherlands Organization for Scientific Research (NWO) for funding the Netherlands Proteomics Center, through the X-omics Road Map program (project 184.034.019), and the EU Horizon 2020 program INFRAIA project Epic-XS (Project 823839). MJvG acknowledges the Amsterdam Infection and Immunity Institute for funding this work through the COVID-19 grant (24175).

## Authors’ contributions

Britt J. van Keulen: 1-4

Michelle Romijn: 1-4

Albert Bondt: 1-4

Kelly Dingess: 1-4

Eva Kontopodi: 1-4

Karlijn van der Straten: 1-4

Maurits A. den Boer: 2-4

Berend J Bosch: 2-4

Philip J.M. Brouwer: 2-4

Christianne J.M. de Groot: 1-4

Max Hoek: 2-4

Wentao Li: 2-4

Dasja Pajkrt: 1-4

Rogier W. Sanders: 2-4

Anne Schoonderwoerd: 2-4

Gestur Vidarsson: 1-4

Koert J. Stittelaar: 1-4

Rian A.H. Timmermans: 1-4

Theo T. Rispens: 1-4

Kasper A. Hettinga: 1-4

Marit J. van Gils: 1-4

Albert J.R. Heck: 1-4

Johannes B. van Goudoever: 1-4

1: Substantial contributions to the conception or design of the work, or the acquisition, analysis or interpretation of data for the work; and

2: Drafting the work or revising it critically for important intellectual content; and

3: Final approval of the version to be published; and

4: Agreement to be accountable for all aspects of the work in ensuring that questions related to the accuracy or integrity of any part of the work are appropriately investigated and resolved.

## Ethics committee approval

Ethical approval was obtained from the Medical Ethics Committee of Amsterdam UMC, location VUmc and written informed consent was obtained from all participants.

## Disclosure

Johannes B. van Goudoever is founder and director of the Dutch National Human Milk Bank and member of the National Health Council. He has been member of the National Breastfeeding Council from 2010-March 2020. The study was funded by de Stichting Steun Emma Kinderziekenhuis.

This research project was registered at the Dutch Trial Register on May 1st, 2020, number: NL 8575

URL: https://www.trialregister.nl/trial/8575

**eFigure 1. Association of normalized antibody levels in breastmilk and serum against S protein of SARS-CoV-2 using ELISA, and sampling time point in weeks after symptom onset for PCR confirmed SARS-CoV-2 infected mothers.** Blue dots and red squares represent IgG levels in serum and IgA levels in milk respectively.

**eFigure 2. Association between IgA levels in breastmilk targeting S protein of SARS-CoV-2 measured by ELISA, and neutralization titers of breastmilk against clinical isolate SARS-CoV-2 virus.** Red dots, blue dots and greens dots represent unpasteurized (UP), High Pressure Pasteurized(HPP) and Holder Pasteurized (HoP) breastmilk respectively.

**eTable 1. Overview of all raw data**.

## References

1. Mullard, A., COVID-19 vaccine development pipeline gears up. Lancet, 2020. 395(10239): p. 1751–1752.

2. Nutrition, E.C.o., et al., Breast-feeding: A commentary by the ESPGHAN Committee on Nutrition. J Pediatr Gastroenterol Nutr, 2009. 49(1): p. 112–25.

3. Chantry, C.J., C.R. Howard, and P. Auinger, Full breastfeeding duration and associated decrease in respiratory tract infection in US children. Pediatrics, 2006. 117(2): p. 425–32.

4. Hanson, L.A., Breastfeeding provides passive and likely long-lasting active immunity. Ann Allergy Asthma Immunol, 1998. 81(6): p. 523–33; quiz 533–4, 537.

5. Mantis, N.J., N. Rol, and B. Corthesy, Secretory IgA’s complex roles in immunity and mucosal homeostasis in the gut. Mucosal Immunol, 2011. 4(6): p. 603–11.

6. Van de Perre, P., Transfer of antibody via mother’s milk. Vaccine, 2003. 21(24): p. 3374–6.

7. Hutchings, A.B., et al., Secretory immunoglobulin A antibodies against the sigma1 outer capsid protein of reovirus type 1 Lang prevent infection of mouse Peyer’s patches. J Virol, 2004. 78(2): p. 947–57.

8. Kang, S., et al., Crystal structure of SARS-CoV-2 nucleocapsid protein RNA binding domain reveals potential unique drug targeting sites. Acta Pharm Sin B, 2020.

9. Schlaudecker, E.P., et al., IgA and neutralizing antibodies to influenza a virus in human milk: a randomized trial of antenatal influenza immunization. PLoS One, 2013. 8(8): p. e70867.

10. Demers-Mathieu, V., et al., Antenatal Influenza A-Specific IgA, IgM, and IgG Antibodies in Mother’s Own Breast Milk and Donor Breast Milk, and Gastric Contents and Stools from Preterm Infants. Nutrients, 2019. 11(7).

11. Fox, A., et al., Evidence of a significant secretory-IgA-dominant SARS-CoV-2 immune response in human milk following recovery from COVID-19. medRxiv, 2020: p. 2020.05.04.20089995.

12. Italian Association of Human Milk Banks Associazione Italiana Banche del Latte Umano, D., et al., Guidelines for the establishment and operation of a donor human milk bank. J Matern Fetal Neonatal Med, 2010. 23 Suppl 2: p. 1–20.

13. Ford, J.E., et al., Influence of the heat treatment of human milk on some of its protective constituents. J Pediatr, 1977. 90(1): p. 29–35.

14. Demers-Mathieu, V., et al., Differences in Maternal Immunoglobulins within Mother’s Own Breast Milk and Donor Breast Milk and across Digestion in Preterm Infants. Nutrients, 2019. 11(4).

15. Viazis, S., B.E. Farkas, and J.C. Allen, Effects of high-pressure processing on immunoglobulin A and lysozyme activity in human milk. Journal of Human Lactation, 2007. 23(3): p. 253–261.

16. Wesolowska, A., et al., Innovative Techniques of Processing Human Milk to Preserve Key Components. Nutrients, 2019. 11(5).

17. Wrapp, D., et al., Cryo-EM Structure of the 2019-nCoV Spike in the Prefusion Conformation. bioRxiv, 2020.

18. Vogelzang, E.H., et al., Development of a SARS-CoV-2 total antibody assay and the dynamics of antibody response over time in hospitalized and non-hospitalized patients with COVID-19. medRxiv, 2020: p. 2020.06.17.20133793.

19. Martinez-Monteagudo and V.M. Balasubramaniam, Fundamentals and Applications of High-Pressure Processing Technology. Food Engineering Series, ed. V.M. Balasubramaniam, G.V. Barbosa-Canovas, and H. Lelievel. 2016, High Pressure Processing of Food: Springer, New York, NY.

20. Timmermans, R., et al., Comparing thermal inactivation to a combined process of moderate heat and high pressure: Effect on ascospores in strawberry puree. Int J Food Microbiol, 2020. 325: p. 108629.

21. Okba, N.M.A., et al., SARS-CoV-2 specific antibody responses in COVID-19 patients. medRxiv, 2020: p. 2020.03.18.20038059.

22. Sterlin, D., et al., IgA dominates the early neutralizing antibody response to SARS-CoV-2. medRxiv, 2020: p. 2020.06.10.20126532.

23. Sousa, S.G., I. Delgadillo, and J.A. Saraiva, Effect of thermal pasteurisation and high-pressure processing on immunoglobulin content and lysozyme and lactoperoxidase activity in human colostrum. Food Chemistry, 2014. 151: p. 79–85.

24. Viazis, S., B.E. Farkas, and L.A. Jaykus, Inactivation of bacterial pathogens in human milk by high-pressure processing. Journal of Food Protection, 2008. 71(1): p. 109–118.

25. Peila, C., et al., The Effect of Holder Pasteurization on Nutrients and Biologically-Active Components in Donor Human Milk: A Review. Nutrients, 2016. 8(8).

26. Stringhini, S., et al., Seroprevalence of anti-SARS-CoV-2 IgG antibodies in Geneva, Switzerland (SEROCoV-POP): a population-based study. Lancet, 2020.

